# The impact of urban vs rural environments on driving in ageing

**DOI:** 10.1101/2024.08.12.24310574

**Authors:** Sol Morrissey, Stephen Jeffs, Rachel Gillings, Mizanur Khondoker, Mary Fisher-Morris, Ed Manley, Michael Hornberger

## Abstract

**Background and objectives:** Older rural drivers are more dependent on driving than urban drivers to maintain community mobility due to reduced availability of transportation alternatives. Yet it is not understood how cognition impacts driving mobility and road safety across urban vs rural settings. The present study therefore aimed to establish whether cognitive changes impacted driving mobility and road safety differently across rural and urban older drivers.

**Research Design and Methods:** 969 older drivers (mean age: 71.01) were recruited for a prospective cohort study. Participants completed self-reported driving behaviour and road traffic incident (RTI) history questionnaires before completing an objective cognitive testing battery to establish global cognitive functioning; and were invited back to repeat the study procedure one-year later.

**Results:** We find that older rural drivers have a greater driving mobility than older urban drivers and are less likely to reduce their driving mobility over time, as only urban residents with cognitive decline reduced their driving space. We further corroborate previous findings that RTI incidence is greater within urban areas and establish a distinct association between worse cognitive functioning and RTI risk solely in urban residents.

**Discussion and implications:** Overall, we show for the first time how the interaction of age-related cognitive changes with geographical settings impact driving mobility and road safety in urban and rural areas. This paves the way for informed policymaking and future research directions to navigate driving cessation and improved road safety in ageing.

## Introduction

Driving mobility is vital for maintaining independence amongst older adults (Eby & Molnar, 2009). This is particularly true for individuals living in rural settings, where reduced public transportation access and greater distances to amenities require greater reliance on personal vehicles for daily activities and social engagements (Arcury et al., 2005; Hamano et al., 2016). However, older adults typically self-regulate and reduce their driving mobility as they age, due to the ageing process (Oxley et al., 2010; Payyanadan et al., 2018). With the demographic shift towards an ageing population, and the increase of older adults living in rural areas within Western countries, it is important to understand how changes to cognitive functioning interact with geographical settings to inform how older adults can continue to meet their driving transportation needs whilst maintaining road safety.

Within the UK, the majority of older adults live in rural areas (Office for National Statistics, 2024). Rural drivers have previously been found to depend substantially more on personal vehicle transportation as their only method of transportation compared to those who live in cities or small towns (Ritter et al., 2002). This is largely due to rural areas requiring greater travel distances to reach healthcare services, amenities, and resources; as well as having less access to alternate transportation methods. Within the UK, public transportation has been found to be largely unavailable, unreliable, or deficient in rural areas (Jo et al., 2021), increasing the greater reliance on driving for rural residents. It is therefore of little surprise that driving is regarded more important to individuals living within rural areas (Strogatz et al., 2020), who typically report travelling further distances than urban residents (Payyanadan et al., 2018; Pucher & Renne, 2005).

Despite greater reliance of driving in rural areas, there is a greater risk of fatalities on rural roads, as US-based studies have shown that older adults are over two-times more likely to have fatal road traffic incidents on rural roads than urban roads (Zwerling et al., 2005). Indeed, government statistics show that although urban roads amount in a greater likelihood of overall road traffic incidents (RTIs), largely because of a more dynamic road traffic environment, rural roads are a greater risk for fatal RTIs (Department for Transport, 2023). This is due to less safe aspects of the road environment in rural areas, including narrow roads and higher road speeds (Payyanadan et al., 2018; Thompson et al., 2013). Additionally, it may be that greater dependency on driving in rural areas means that older drivers in these areas may be less likely to self-regulate their driving despite cognitive impairments reducing their driving safety (Byles & Gallienne, 2012; Hanson & Hildebrand, 2011).

During the ageing process, cognitive changes are associated with reduced driving safety (Depestele et al., 2020; Stefanidis et al., 2023). Older drivers typically compensate for these changes by self-regulating their driving, adapting when and where they drive to maintain road safety (Devlin & McGillivray, 2014). Although previous research has been conducted on the interaction of physical impairments on driving mobility within rural and urban environments, showing that measures of physical functioning were more predictive of driving behaviour in larger urban cities (Anstey et al., 2005; O’Connor et al., 2012), research has not yet established how cognitive functioning is associated with driving changes across rural and urban areas. Our research group previously established that spatial orientation is the signature cognitive marker for driving frequency and difficulty in older age (Morrissey, Jeffs, Gillings, Khondoker, Patel, et al., 2024), and that use of GPS technology can ameliorate cognitive impairments to increase driving mobility (Morrissey et al., 2024). However, it is not yet understood how cognitive impairments may interact with driving mobility and safety across geographical settings. This is important to establish, as individuals who cease driving due to self-regulation from cognitive impairments in rural areas often have less alternate transportation methods to maintain social mobility, and those who do not self-regulate effectively may be at greater risk of RTIs.

The current study addresses these gaps in knowledge by establishing how driving mobility changes across rural and urban settings over a one-year period within a large sample of community-dwelling older adult drivers. We will further establish how road safety differs across rural and urban environments. Finally, we will explore how cognitive changes over one-year are associated with changes in driving mobility and driving safety across geographical settings. Specifically, we will i) compare driving characteristics and mobility across geographical settings; ii) assess how road traffic incident frequency interacts with cognitive functioning across geographical settings; iii) examine how driving mobility changes over time across geographical settings; and iv) identify whether global cognitive changes are associated with changes to driving mobility within rural and urban areas separately. We hypothesise that i) drivers within rural areas will rely more upon driving their personal vehicles than community transportation or public transport; ii) drivers in rural areas will demonstrate greater driving frequency and space than individuals in urban areas, as they will be more dependent on driving to meet their mobility needs; iii) drivers in urban environments will experience more road traffic incidents due to driving more frequently in more dynamic, high-traffic environments; iv) urban older drivers will show a reduced driving mobility over time, whereas this is maintained in rural older drivers; and v) older drivers with global cognitive changes living in urban areas will show greater reduction in their driving mobility compared to those living in rural areas.

## Methods

### Participant recruitment

969 older adults (mean age: 71.01, 540 female, rural: 296) were recruited between February 2021 and August 2021 to complete the study. The inclusion criteria for the study were being age 65 or older, holding a valid driving license, and being a regular driver (driving at least once per week). The exclusion criteria for the study were not driving regularly, having a medical condition that contraindicates driving, having an untreated significant visual or physical impairment, having a diagnosis of mild cognitive impairment or dementia, taking medications for dementia, and high alcohol consumption (> 45 units per week). Participants were recruited via online and media advertisement. Signed informed consent was obtained from each participant prior to conducting the experimental protocol and data was attributed anonymously. Ethical approval for the study was provided by the Faculty of Medicine and Health Sciences Research Ethics Committee at the University of East Anglia (FMH2019/20-134).

### Procedure

Participants initially completed online questionnaires related to their demographic information, driving habits, health status, driving history, driving habits, and a custom driving-based navigation questionnaire. Following this, participants completed a neuropsychological testing battery assessing cognitive performance across a variety of domains, including reaction speed, processing speed, executive functioning, spatial working memory, episodic memory, visuospatial functioning, and spatial orientation (see Morrissey et al., 2024). Participants were then invited to complete a follow-up testing phase one year after baseline data collection, undergoing the same procedure. 574 participants took part in the follow-up testing phase (mean age: 71.95, 314 female, 174 rural).

### Driving mobility and safety measures

Driving mobility and safety measures were derived from the Driving Habits Questionnaire (DHQ), as well as novel Driving History and Road Traffic Incident (RTI) questionnaires. Driving mobility measures included annual mileage, weekly driving days, driving space (the geographical area in which people drive), weekly trips, maximum weekly trip distance, situation avoidance, driving speed (relative to the general flow of traffic), and transport reference (Drive yourself, Driven by someone else, Public transport). Driving safety was measured by whether someone was in a recent RTI (within the past 3 years). We also collected the number of in-vehicle technologies used (parking assistance, cruise control, lane control, sat-nav, and Bluetooth) (see Supplementary Table 1 for detailed information on mobility and safety measures).

### Statistical Analysis

Participants were divided into rural or urban groups depending on the outward code (the first part) of their postcode location based on the 2011 Rural-Urban classification data (Department for Environment, Food & Rural Affairs, 2021). Differences in driving characteristics between people living in rural and urban areas were established using two sample t-tests and chi-squared tests for continuous and categorical variables respectively. Analyses of Covariances (ANCOVAs) were conducted to assess whether driving mobility differed across environmental locations after controlling for age as a covariate. In assessing how avoidance of driving situations differed across environmental locations, weekly driving days was added to the model as a covariate. A Pearson’s chi-square test was conducted to establish whether there were differences in transport preferences (Drive yourself, Someone else drives, or Public transport/ Taxi) across environmental locations. A binary logistic regression was used to assess whether environmental location predicted whether individuals were more likely to have a recent road traffic incident after accounting for age and annual mileage as covariates as they have previously been associated with increased road traffic incident risk. Post-hoc logistic regression analyses were then conducted to assess whether global cognitive functioning was associated with recent RTIs between rural and urban environments separately after controlling for age and mileage. Individual spatial orientation tests were not assessed with recent RTIs due to few rural residents with a recent RTI completing spatial orientation tests. A post-hoc independent samples t-test analysis was then conducted to assess whether the annual mileage for individuals who had experienced a recent RTI differed across rural and urban residents. We then assessed whether driving mobility changes over a one-year period were associated with environmental location using linear mixed effect (LME) modelling. For LME analysis, difference in driving mobility was calculated by subtracting the baseline score from the follow-up score. Age was included as a covariate and a random intercept term was added to the model to account for individual variability. We then assessed whether global cognitive performance was associated with driving mobility variables using linear regression models across geographical settings, separately. Cognitive functioning across both geographical settings was comparable as a Mann-Whitney U test revealed that there was no significant difference in global cognitive performance between rural and urban areas (W = 39425, *p* = 0.14). Following this, we assessed whether cognitive change over time was associated with change in driving mobility within environmental locations separately. To develop a global cognitive change score, cognitive data (reaction time, processing speed, executive functioning, spatial working memory, episodic memory) was standardised within each cognitive measure using the grand mean from both timepoints, and average performance across all tasks was derived across baseline and follow-up test phases. Cognitive change was established by subtracting follow-up global cognition from baseline global cognition. Spatial orientation tests (allocentric & egocentric orientation) were omitted for global cognitive change measurement as fewer participants completed these tests across both testing phases and therefore there would have been a substantive reduction in global cognitive change data (172 compared to 311 participants). Post-hoc analysis was therefore conducted to establish whether spatial orientation performance change over time was associated with change in driving mobility changes across environmental locations separately.

To account for potential measurement error of online testing, outliers were assessed for baseline and follow-up data using boxplots, Q-Q plots, and histograms. For online cognitive data, extreme outliers outside of 3 *SD* were removed for reaction time (baseline: 8, follow-up: 6), trail-making test - A(10, 6), trail-making test – B (16, 8), spatial working memory (5, 0), allocentric orientation (2, 0), egocentric orientation (2, 0), and subjective sense of direction (5, 3). Extreme values above and below the 99^th^ percentile were removed for recognition memory (8, 5) and source memory (8, 5). For self-reported driving data, extreme outliers were also removed for typical annual mileage (18), driving space (1, 0), weekly trips (13, 2), and weekly trip distance (11, 12), number of passengers (7), years spent with current car (8), and cars regularly driven (8). Weekly trips and maximum weekly trip distance variables were given a logarithmic transformation for analysis due to high positive skewness. For ANCOVA and LME analysis, checking normality of outcome variables was conducted using visual inspection of histograms and normality of residuals was conducted by QQ-Plots. Linearity assumptions and multicollinearity were checked for regression analyses. A significance threshold of 0.05 was used to assess statistical significance. All analysis was carried out in R (version 4.3.1) using car, lme4, and nlme packages.

## Results

### Driving characteristics of older rural and urban residents in the UK

Within our cohort, individuals living in rural environments had more years of driving experience (*p*<.05), and less use of in-vehicle technology than urban drivers (*p*<.05) (see Table 1). 125 participants self-reported recent RTIs (95 living in urban locations).

**Table 1.** Participant demographic and driving characteristics.

| **Table 1. Participant demographic and driving characteristics** |  |  |  |  |
| --- | --- | --- | --- | --- |
| **Variable** | **Environment** |  | ***p*-value** | **Effect size (*d*)** |
|  | **Rural** | **Urban** |  |  |
| Participants | 296 | 673 |  |  |
| Age (years) | 71.38 (5.30) | 70.85 (4.78) | 0.14 | 0.11 |
| Gender (% female) | 52.36 | 57.21 | 0.18 | 0.94 |
| Education (years) | 14.78 (2.85) | 14.90 (2.71) | 0.54 | 0.04 |
| Driving experience (years) | 50.27 (7.13) | 48.96 (7.52) | 0.01 | 0.18 |
| Subjective driving ability | 3.79 (0.62) | 3.79 (0.65) | 0.99 | 0.00 |
| Cars regularly driven | 1.34 (0.57) | 1.31 (0.51) | 0.45 | 0.06 |
| Time with current vehicle (years) | 2.82 (3.45) | 3.23 (3.80) | 0.11 | 0.11 |
| N. of regular passengers | 1.07 (1.11) | 1.18 (1.19) | 0.20 | 0.09 |
| Use of in-vehicle technology | 0.91 (0.69) | 1.03 (0.81) | 0.02 | 0.16 |
| Sat-Nav use | 1.01 (1.02) | 1.18 (1.04) | 0.14 | 0.16 |
| Driving speed | 3.03 (0.49) | 2.99 (0.43) | 0.18 | 0.10 |
| Note.<br><sup>a</sup> Welch's two sample t test conducted for group differences. Chi squared test of independence used for Gender analysis.<br><sup>b</sup> Cramér's V effect size used for Gender analysis. Cohen's D effect sizes calculated for other variables. |  |  |  |  |

### Impact of urban vs rural environment on driving mobility

Rural residents showed a significantly greater driving space (*F*(1, 939) = 6.164, *p*<.05, η_p_^2^ (partial eta squared) = 0.01); typical annual mileage, (*F*(1, 924) = 23.684, p < .001, η_p_^2^ = 0.02); higher maximum weekly trip distance (*F*(1, 554) = 17.960, *p*<.001, η_p_^2^ = 0.03), but made less weekly driving trips than urban residents (*F*(1, 588) = 5.886, *p* <.05, η_p_^2^ = 0.01) (see Figure 1). Urban residents avoided more driving situations than rural residents (*F*(1, 943) = 9.701, *p*<.01, η_p_^2^ = 0.01. There were no significant differences in driving days or relative driving speed between groups.

**Figure 1.**
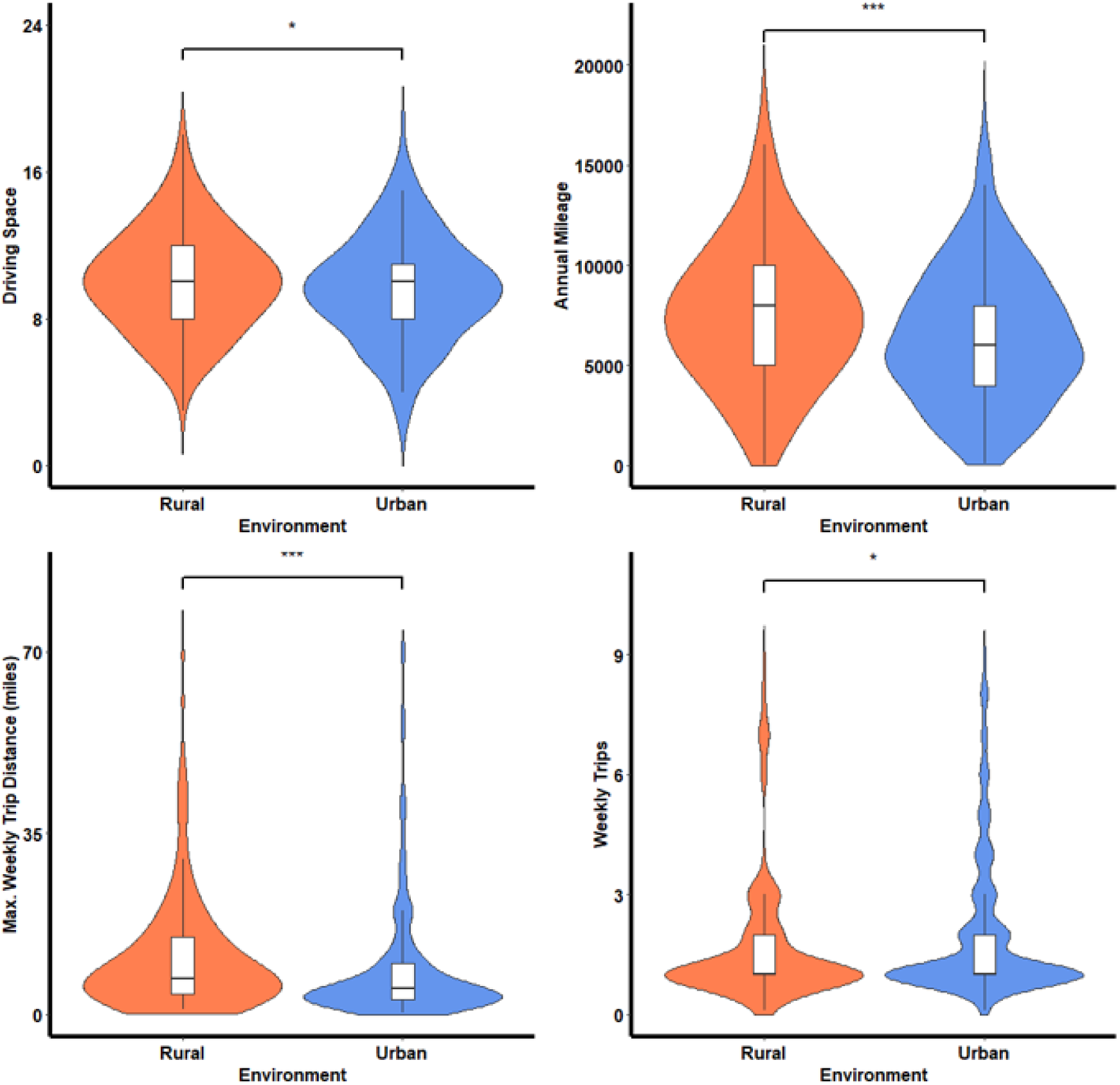
Driving mobility differences across rural and urban settings.

Significant differences in transport preferences were found between rural and urban residents, (χ2 = 7.27, df = 2, *p* < .05), with rural residents less likely to use public transport or rely upon a friend to drive them than people living in urban areas.

### Impact of urban vs rural environment on driving safety

Urban residents were more likely to have been in a recent road traffic incident than rural residents (OR = 1.57, *p*<.05, CI[1.02, 2.48]) (see Figure 2). Worse global cognitive functioning was predictive of a greater incidence of RTIs within urban residents (OR = 1.98, *p*<.05, CI[1.00, 3.88]), but not rural residents (see Table 2). Among individuals involved in a recent RTI, there was no significant difference in typical annual mileage between rural and urban residents.

**Figure 2.**
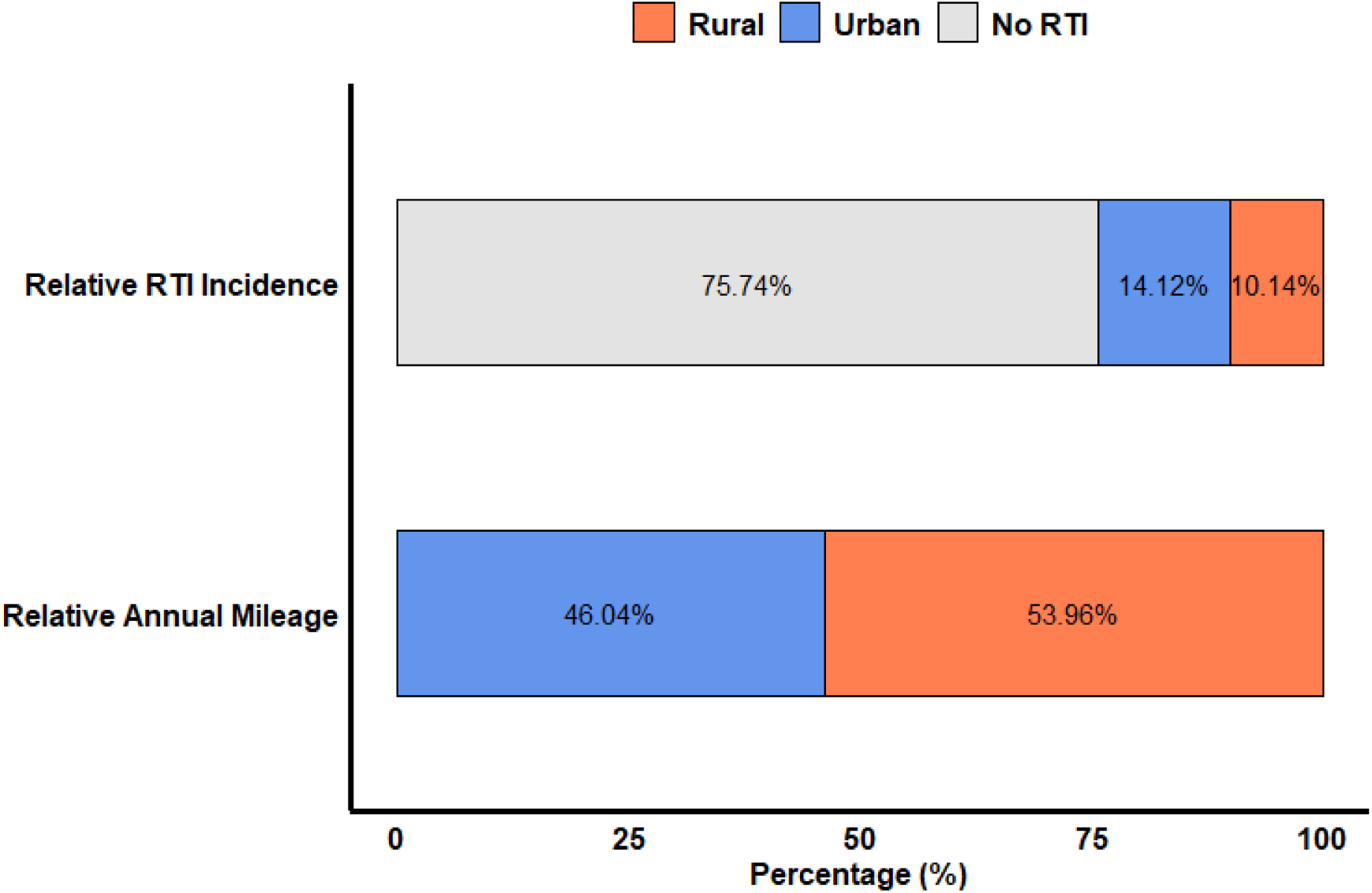
Relative road traffic incident incidence and relative annual mileage across rural and urban areas.

**Table 2.**
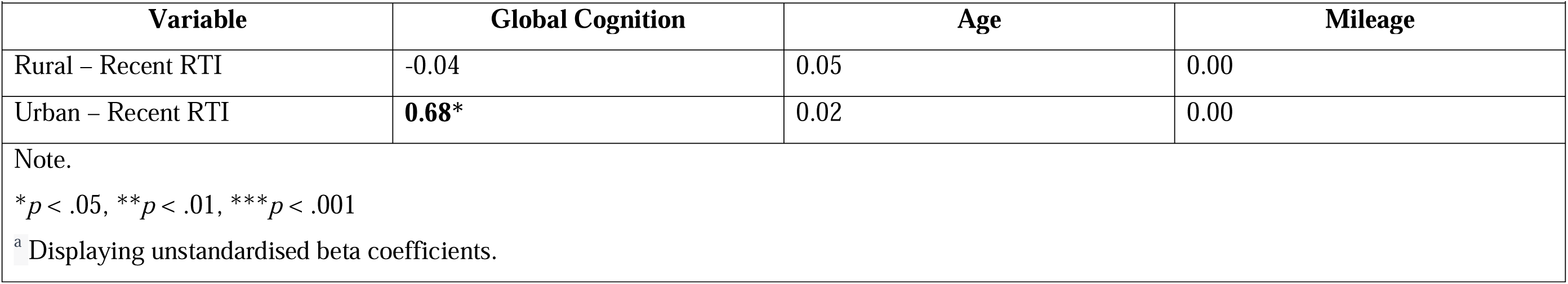
Multiple logistic regression analysis comparing recent road traffic incident (RTI) occurrence across rural and urban environments.

|  |  |  |  |
| --- | --- | --- | --- |
| Variable | Global Cognition | Age | Mileage |
| Rural – Recent RTI | -0.04 | 0.05 | 0.00 |
| Urban – Recent RTI | **0.68\*** | 0.02 | 0.00 |
| Note.<br>$*p < .05$, $**p < .01$, $***p < .001$<br><sup>a</sup> Displaying unstandardised beta coefficients. | | | |

### Impact of cognitive performance across urban vs. rural environments

Worse global cognitive functioning was associated with a smaller driving space (β = -1.12, *p*<.05, CI[-2.04, -0.20]) and slower driving speed (β = -0.22, *p*<.05, CI[-0.39, -0.05]) among rural residents, and less annual mileage amongst urban residents (β = -803.09, *p*<.05, CI[-1581.20, -24.98]). Post-hoc spatial orientation tests revealed that worse allocentric orientation was associated with less annual mileage (β = -596.41, *p*<.001, CI[-943.17, -249.66]) and smaller driving space (β = -0.361, *p*<.01, CI[-0.62, -0.10]) within rural areas, and greater avoidance of driving situations (β = 0.115, *p*<.01, CI[0.03, 0.20]) within urban residents. Worse egocentric orientation performance was associated with reduced driving space (β = - 0.01, *p*<.05, CI[-0.02, -0.00]) and greater avoidance of driving situations (β = 0.006, *p*<.01, CI[0.00, 0.01]) in urban residents (see Table 3).

**Table 3.**
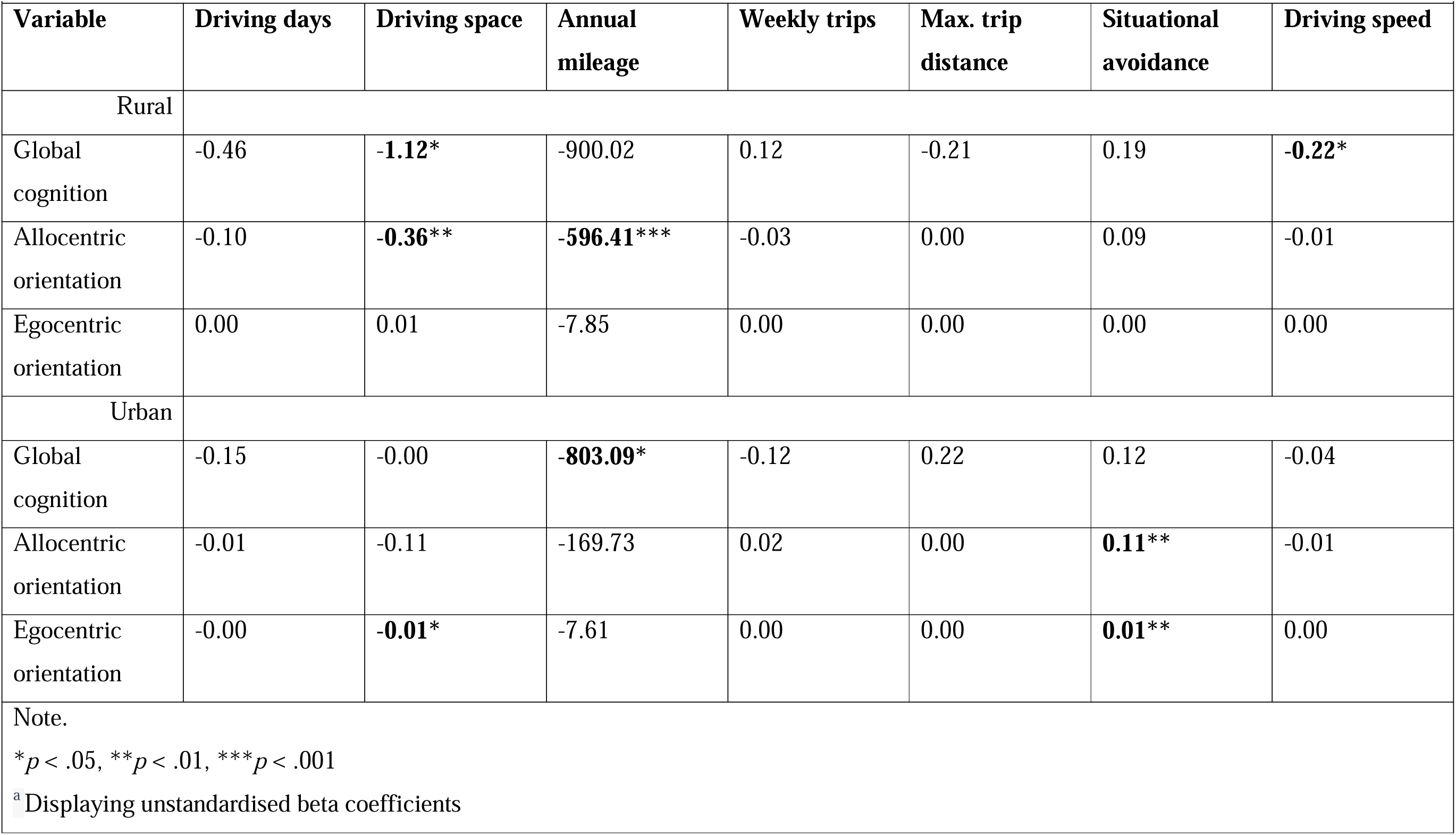
Multiple linear regression analysis establishing how cognitive performance interacts with driving mobility across rural and urban environments.

| Variable | Driving days | Driving space | Annual mileage | Weekly trips | Max. trip distance | Situational avoidance | Driving speed |
| --- | --- | --- | --- | --- | --- | --- | --- |
| Rural |  |  |  |  |  |  |  |
| Global cognition | -0.46 | **-1.12\*** | -900.02 | 0.12 | -0.21 | 0.19 | **-0.22\*** |
| Allocentric orientation | -0.10 | **-0.36\*\*** | **-596.41\*\*\*** | -0.03 | 0.00 | 0.09 | -0.01 |
| Egocentric orientation | 0.00 | 0.01 | -7.85 | 0.00 | 0.00 | 0.00 | 0.00 |
| Urban |  |  |  |  |  |  |  |
| Global cognition | -0.15 | -0.00 | **-803.09\*** | -0.12 | 0.22 | 0.12 | -0.04 |
| Allocentric orientation | -0.01 | -0.11 | -169.73 | 0.02 | 0.00 | **0.11\*\*** | -0.01 |
| Egocentric orientation | -0.00 | **-0.01\*** | -7.61 | 0.00 | 0.00 | **0.01\*\*** | 0.00 |

Note.
\* $p < .05$ , \*\* $p < .01$ , \*\*\* $p < .001$
<sup>a</sup> Displaying unstandardised beta coefficients

### Longitudinal driving changes across urban vs rural environments

Urban residents exhibited a greater decline in their driving space over time (β = -0.652, *p*<.01, CI[-1.10, -0.21]), and were more likely to avoid more driving behaviours over time than rural residents (β = 0.334, *p*<.001, CI[0.138, 0.530]). No significant differences were found in driving days, weekly trips, maximum weekly trip distance, or driving speed (see Supplementary Table 2).

No significant associations were found between global cognitive changes and driving mobility over time across environmental location. Post-hoc analysis of the association between spatial orientation performance and driving mobility across rural and urban locations showed that in urban residents the decline in allocentric orientation performance predicted reduced driving space over time (β = 0.338, *p*<.05, CI[0.02, 0.65]).

## Discussion

Within a large sample of healthy older adults, the present study examined how driving mobility and safety differs across rural and urban environments over a one-year period and establishes how this relates to cognitive functioning. Overall, we found that rural residents show a greater driving mobility than urban residents and were less likely to decrease their driving mobility over time. We also demonstrate that worse cognitive performance is associated with lower driving mobility in both rural and urban areas, but only urban residents with decline in spatial orientation ability reduced their driving space over time. Importantly, we corroborate previous findings showing that urban residents were more likely to be in a recent collision than rural residents and build upon previous findings to show that people with worse global cognition are more likely to be in RTIs within urban areas.

Within our sample, approximately 14% of urban residents and 10% of rural residents self-reported a recent RTI, supporting previous evidence that RTIs are more common in urban environments (Merlin et al., 2020). Worse cognitive functioning has previously been associated with an increased presence of RTIs within older age (Ball et al., 2006; Emerson et al., 2012; Fraade-Blanar et al., 2018; Kosuge et al., 2017), however this study is the first to our knowledge to show that worse cognitive functioning is associated with increased RTI risk amongst urban but not rural residents. Urban road environments present greater hazards due to a more dynamic road environment, and cognitive deficits in healthy ageing have previously been associated with experiencing challenges for road features common in urban road environments, such as intersections and higher traffic volume (Morrissey et al., 2024; Swain, McGwin, Antin, Wood, Owsley, 2021; Son, Lee, & Kim, 2011). The heightened risk of RTIs among urban residents may therefore be attributed in part to the interaction between cognitive decline in ageing individuals and the complexities of navigating urban road environments. One potential explanation for the lack of a concurrent effect in rural drivers could be attributed to our observation that rural drivers with worse cognition were more likely to reduce their speed relative to other drivers on the road, but not urban drivers. This differential response may be linked to the perception that altering speed limits poses a greater risk on urban roads compared to rural ones (Cox et al., 2017), possibly due to greater environmental complexity on urban roads requiring more attentional resources. The higher speed limit and less congested nature of rural roads may consequently afford for cognitively impaired rural drivers to compensate by reducing their travel speed, mitigating the risk of RTI involvement. Rural drivers with cognitive impairments who do not reduce their relative speed may therefore be at a greater risk of RTIs, which at higher road speeds are more likely to be fatal. Future work looking more granularly at risky driving behaviour, via sharp decelerating/braking events, may be able to entangle the relationship more accurately between cognitive impairment and driving safety in rural areas.

Aligning with our hypotheses, rural residents demonstrated a greater driving mobility than urban residents: driving at a greater annual mileage, covering greater driving space, and having a higher distance in weekly trips. In reverse, urban residents reported a greater number of weekly trips. The greater reliance on driving in rural areas is consistent with previous US and Australia based findings that older rural drivers show greater mobility than the urban population (Pucher & Renne, 2005; Payyanandan et al., 2018; Byles & Galliene, 2012). Differences found in weekly trip frequency across geographical settings may be related to accessibility of amenities and local services, as urban households living closer to intended destinations would be more likely to take shorter, more frequent trips than more isolated rural residents, who may be less inclined to be on the road again after travelling further distances to reach their destination and may conduct multiple stops in one trip. We also establish that urban residents are more likely to avoid challenging situations than rural residents, corroborating previous focus-group findings where older urban drivers reported greater difficulties in driving through heavy traffic, and preferred using interstate highways as they reduced challenging driving situations (Payyanandan et al., 2018). Therefore, driving in urban areas may present greater possibilities of compensating by avoiding difficult situations, which may not be possible in rural areas where there are fewer route options due to less street network intersections. This is supported by our longitudinal findings, showing that urban residents were more likely to decrease their number of challenging driving situations faced and their driving space after a one-year period compared to rural residents.

The greater reluctance of rural residents to reduce their driving mobility over time may be related to a greater reliance on driving as a transportation method to meet their mobility needs. Rural residents were less likely to rely upon alternate forms of transportation than urban residents, including public transport or relying upon a friend to drive them. Therefore, whilst community transportation is common amongst older adult populations (Kerschner & Rousseau, 2008; Davey, 2006), it may be that this is less prevalent within rural areas and potentially a less viable transportation alternative. Among our sample, however, we found no significant differences in the number of regular driving passengers for rural and urban drivers, indicating that despite potential disparities in transportation options, both rural and urban residents maintain similar levels of social engagement and support through shared mobility experiences.

Within both rural and urban areas, we observe that worse global cognitive functioning was associated with reduced driving mobility. Longitudinally, however, only urban residents with declining allocentric spatial orientation ability reduced their driving mobility, showing a smaller driving space over time. Rural residents with cognitive impairments may therefore be less inclined to reduce their driving than urban residents, possibly due to fewer transportation alternatives to meet their mobility needs. There is a potential bidirectional component to the relationship between allocentric orientation decline and reduced driving space, as it is unclear whether individuals may show reduced driving space because of cognitive decline, or whether individuals are experiencing cognitive decline due to reduced hippocampal activation involved in allocentric spatial processing. Successful allocentric spatial orientation is highly dependent on cognitive mapping within the medial temporal lobe, which is one of the earliest brain areas to undergo neurophysiological changes in advanced normative ageing (Raz et al., 2004). It is possible that due to being more closely located to amenities and services, older urban residents travel less frequently to distant locations over time and engage less with hippocampal-based cognitive mapping processes, reducing their allocentric spatial orientation ability. Maintaining driving in older age and living in more spatially complex environments has previously been associated with reductions in hippocampal brain atrophy in older age (Shimada et al., 2023; Shin et al., 2024). Reducing one’s driving space, and keeping to familiar routes, may therefore result in declining allocentric spatial orientation performance over time due to hippocampal atrophy. Furthermore, as allocentric spatial orientation was the only cognitive modality associated with reductions in driving mobility, this is supportive of previous work showing that allocentric orientation is a key cognitive marker toward driving changes in healthy ageing (Morrissey et al., 2024).

This study provided valuable insights into the interaction between cognition and environment on driving mobility and safety that have several important implications for policymakers and future investigation. Environmentally tailored interventions may be needed to address the specific challenges faced by older drivers in urban and rural settings. For example, urban-focused interventions should emphasise cognitive screening for older drivers and education campaigns on navigating complex urban traffic patterns. Urban design should focus on understanding how cities can support older adults ageing in place and undertake more local activities, as they are more likely to reduce their driving space over time (Vivoda et al., 2017; Wang et al., 2021). In rural areas, interventions should focus on strategies for maintaining mobility and independence while acknowledging the limited availability of alternative transportation options. As rural drivers rely more upon driving to meet their transportation needs, cessation is potentially deeply impactful for their community participation and mobility (Mielenz et al., 2024; Strogatz et al., 2020). Rural communities may therefore benefit from increased support and resources for older adults who face challenges in accessing transportation alternatives. Potential initiatives may include volunteer driver programs, expanded access to public transportation services, and community-based transportation solutions to reduce the impact of driving cessation in older age.

Despite the important findings in our study, there are some limitations. Firstly, in using postcode data to infer urban/rural status, we use between-subject comparisons (alike many driving-environment studies (Dunsire & Baldwin, 1999; Payyanadan et al., 2018; Pucher & Renne, 2005) and do not account for the extent to which individuals drive within rural or urban environments. Future research measuring naturalistic driving can more granularly delineate driving mobility and safety differences across rural and urban environments, establishing how driving mobility changes across the rural-urban scale. Nonetheless, our sample is representable across the UK, as there are approximately 2.5 million older adults living in rural areas and 8 million living in urban areas. Our sample consisted of a similar proportional disparity between rural (296) and urban (673) dwellers. Secondly, as driving mobility and RTI data were self-reported within our study, it is possible that they were prone to inaccuracy and/or bias, as self-report data has been found to differ from objective mobility and crash statistics (McGwin et al., 1998). Using naturalistic driving data to measure driving mobility and objective RTI data provided by crash reports can provide objective and accurate data with which responsibility and cause of the RTI can be ascertained, which will allow for more in-depth analysis on how cognitive functioning interacts with road safety risk. Lastly, the number of self-reported RTIs was low, particularly for rural older adults (30), and did not enable for longitudinal testing of road safety risk. By sampling for participants who had been involved in RTIs in the future, this will enable for a greater number of participants with which to compare to a non-RTI control group.

In conclusion, the present study establishes the differential impact of age-related cognitive changes on driving mobility and safety within rural and urban areas over time, emphasising the importance of considering the interaction between cognitive functioning with regional setting in managing changes to driving safety and mobility in older age. We discuss the implications on maintaining independence in older age, and present future research directions and policymaking options to address the evolving needs of older drivers to promote a safer and sustainable transportation model.

## Funding

This work was supported by the UK Department for Transport (grant number: R208830). This study is supported by the National Institute for Health and Care Research (NIHR) Applied Research Collaboration East of England at Cambridge and Peterborough NHS Foundation Trust. S. Morrissey’s studentship is jointly funded by the Faculty of Medicine & Health Sciences, University of East Anglia (United Kingdom), and the Earle and Stuart Charitable Trust. The views expressed are those of the author(s) and not necessarily those of the NIHR or the Department of Health and Social Care (United Kingdom).

## Data Availability

Data will be made publicly available at publication of the manuscript.

## Acknowledgements

Data used for analysis will be made publicly available upon publication. This study was not pre-registered.

## Supplementary Section

**Supplementary Table 1.**
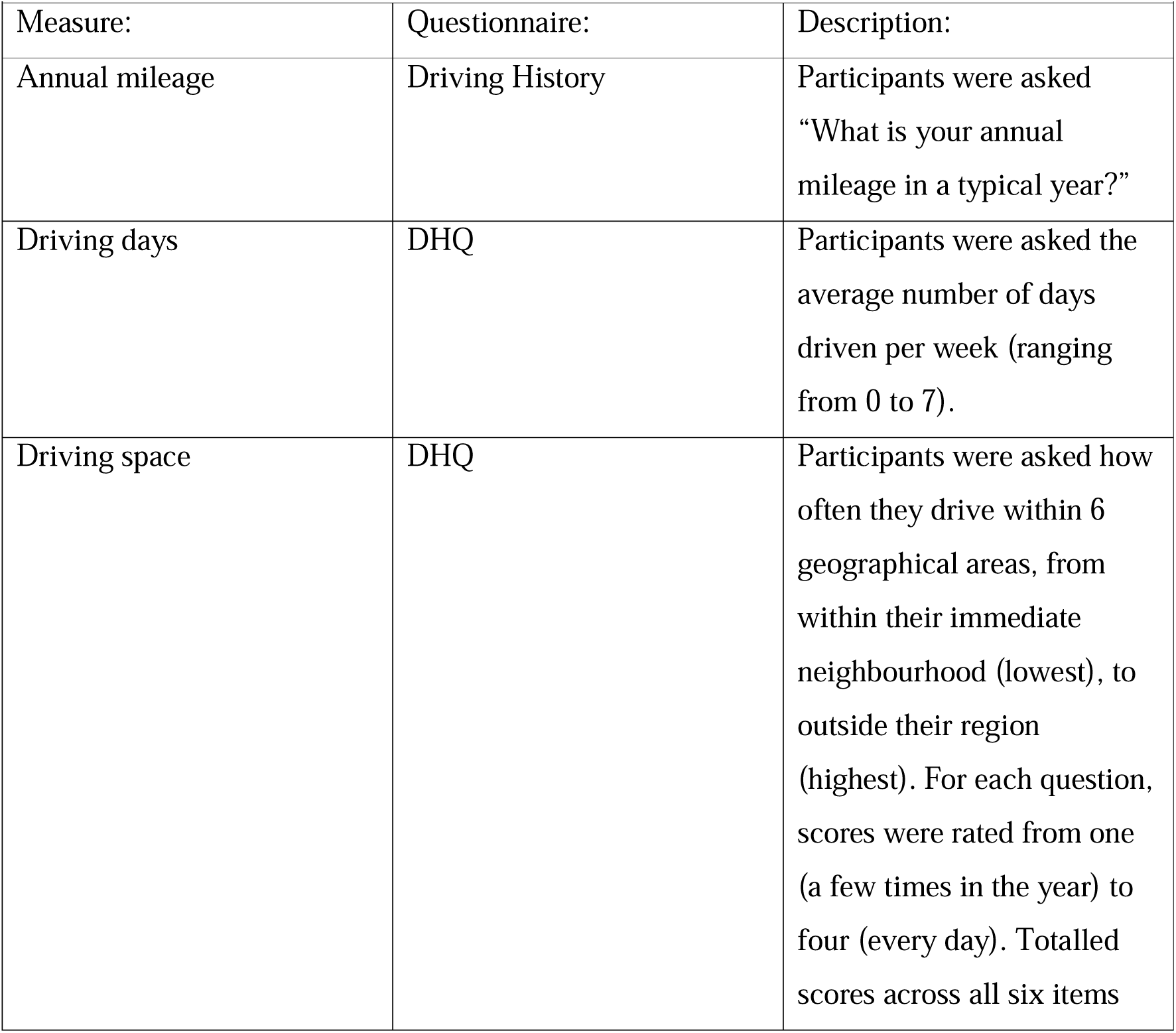
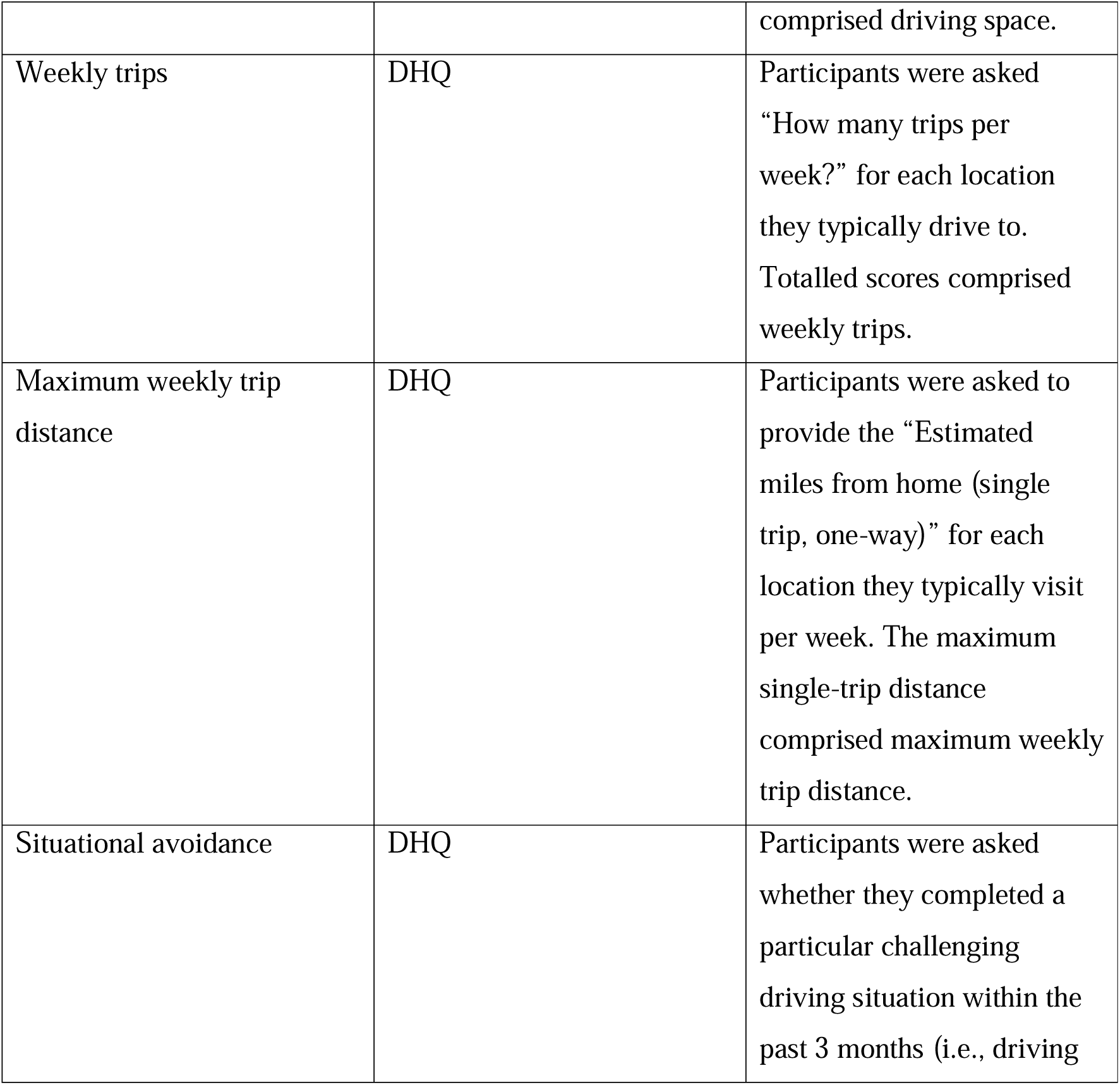
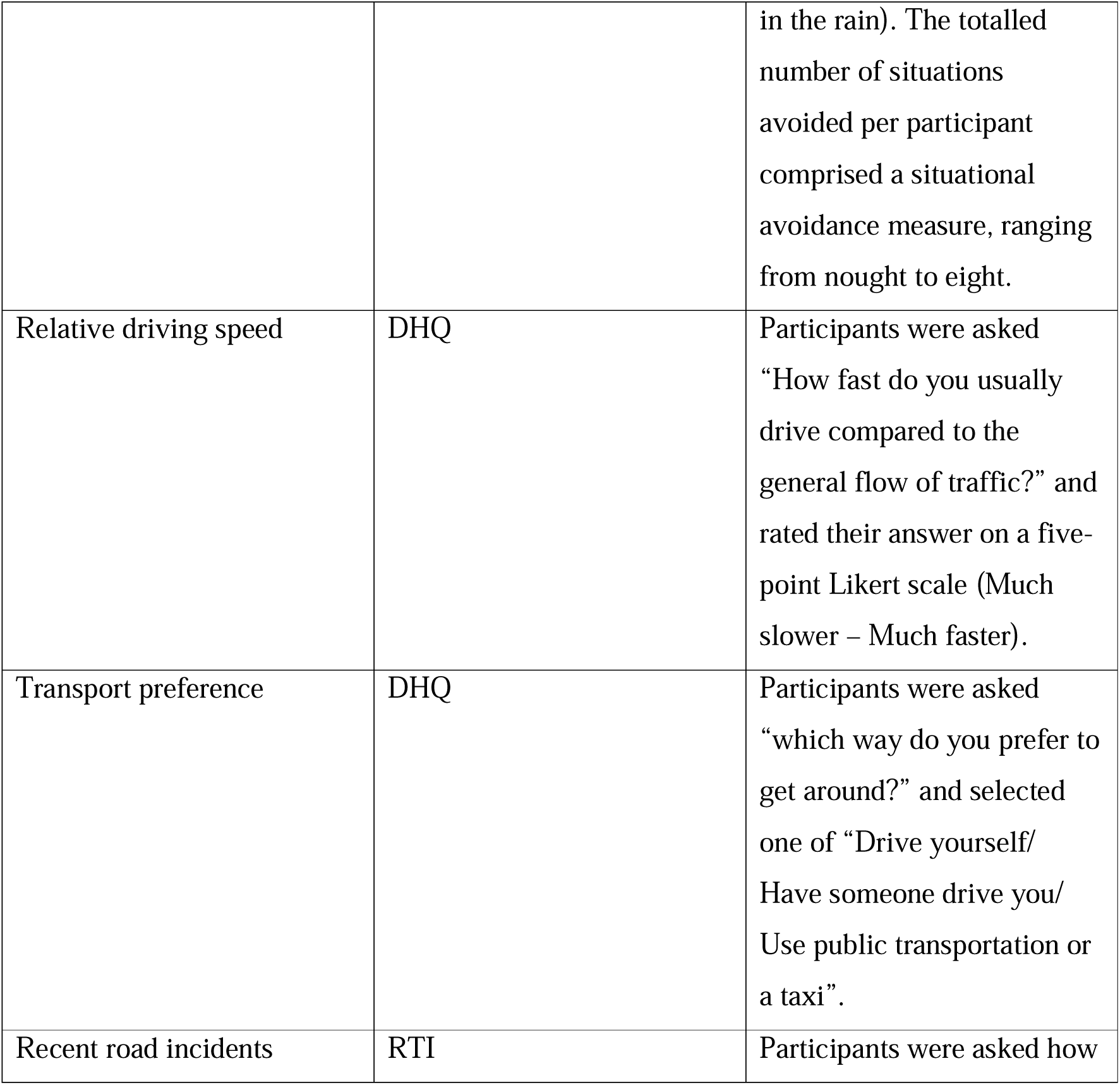
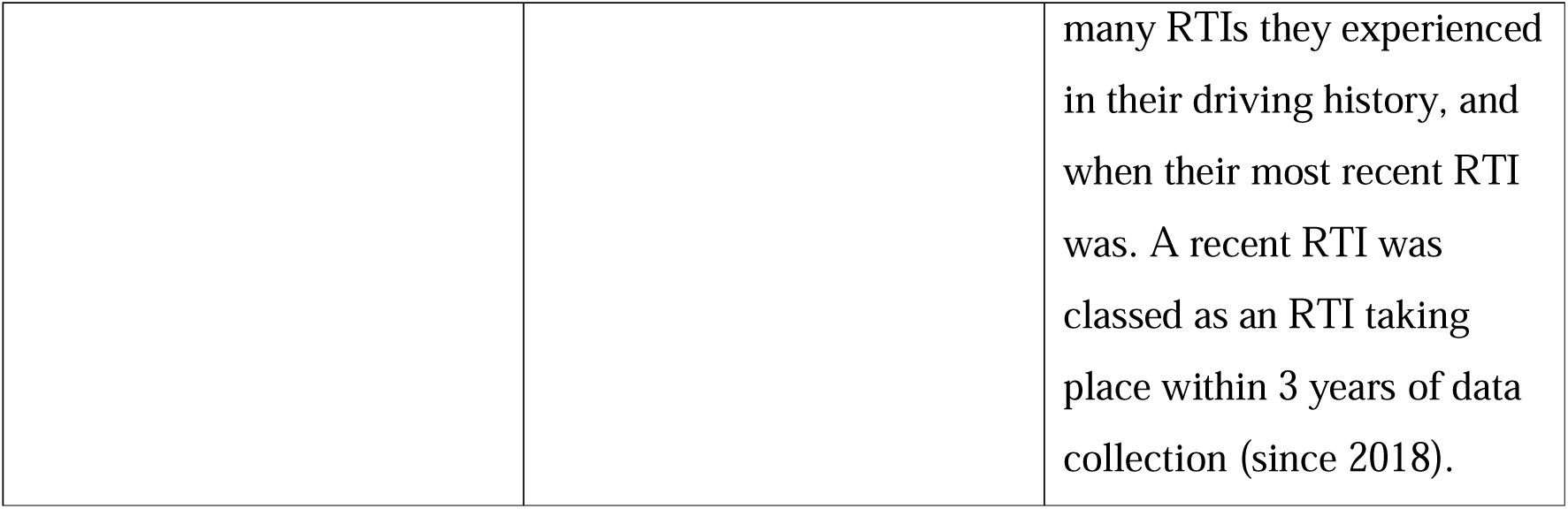
Driving mobility and safety measure descriptions.

**Supplementary Table 2.**
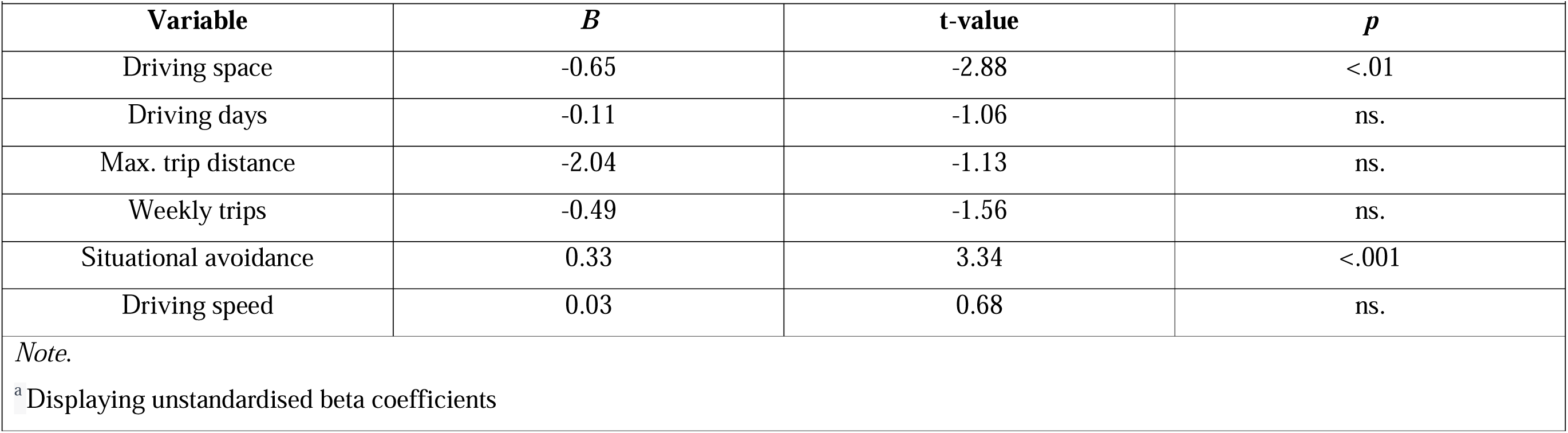
Linear mixed effect model analysis showing how rural and urban environments influence driving mobility.

## Notes

### Competing Interest Statement

The authors have declared no competing interest.

### Funding Statement

This work was funded by the UK Department for Transport (grant number: R208830). This study is supported by the National Institute for Health and Care Research (NIHR) Applied Research Collaboration East of England at Cambridge and Peterborough NHS Foundation Trust. The studentship of S. Morrissey is jointly funded by the Faculty of Medicine & Health Sciences, University of East Anglia (United Kingdom), and the Earle and Stuart Charitable Trust. The views expressed are those of the author(s) and not necessarily those of the NIHR or the Department of Health and Social Care (United Kingdom).

### Author Declarations

Ethical approval for the study was provided by the Faculty of Medicine and Health Sciences Research Ethics Committee at the University of East Anglia (FMH2019/20-134).

## References

Anstey, K. J., Wood, J., Lord, S., & Walker, J. G. (2005). Cognitive, sensory and physical factors enabling driving safety in older adults. Clinical Psychology Review, 25(1), 45–65. 10.1016/J.CPR.2004.07.008

Arcury, T. A., Preisser, J. S., Gesler, W. M., & Powers, J. M. (2005). Access to transportation and health care utilization in a rural region. The Journal of Rural Healthl: Official Journal of the American Rural Health Association and the National Rural Health Care Association, 21(1), 31–38. 10.1111/J.1748-0361.2005.TB00059.X

Ball, K. K., Roenker, D. L., Wadley, V. G., Edwards, J. D., Roth, D. L., McGwin, G., Raleigh, R., Joyce, J. J., Cissell, G. M., & Dube, T. (2006). Can high-risk older drivers be identified through performance-based measures in a Department of Motor Vehicles setting? Journal of the American Geriatrics Society, 54(1), 77–84. 10.1111/J.1532-5415.2005.00568.X

Byles, J., & Gallienne, L. (2012). Driving in older age: a longitudinal study of women in urban, regional, and remote areas and the impact of caregiving. Journal of Women & Aging, 24(2), 113–125. 10.1080/08952841.2012.639661

Cox, J. A., Beanland, V., & Filtness, A. J. (2017). Risk and safety perception on urban and rural roads: Effects of environmental features, driver age and risk sensitivity. Traffic Injury Prevention, 18(7), 703–710. 10.1080/15389588.2017.1296956

Department for Environment, Food & Rural Affairs. (2021). The 2011 Rural-Urban Classification for Output Areas in England

Department for Transport. (2023). Reported road collisions, vehicles and casualties tables for Great Britain - GOV.UK. https://www.gov.uk/government/statistical-data-sets/reported-road-accidents-vehicles-and-casualties-tables-for-great-britain

Depestele, S., Ross, V., Verstraelen, S., Brijs, K., Brijs, T., van Dun, K., & Meesen, R. (2020). The impact of cognitive functioning on driving performance of older persons in comparison to younger age groups: A systematic review. Transportation Research Part F: Traffic Psychology and Behaviour, 73, 433–452. 10.1016/J.TRF.2020.07.009

Devlin, A., & McGillivray, J. A. (2014). Self-regulation of older drivers with cognitive impairment: A systematic review. Australasian Journal on Ageing, 33(2), 74–80. 10.1111/AJAG.12061

Dunsire, M., & Baldwin, S. (1999). Urban-rural comparisons of drink-driving behaviour among late teens: a preliminary investigation. *Alcohol and Alcoholism (Oxford*, Oxfordshire), 34(1), 59–64. 10.1093/ALCALC/34.1.59

Eby, D. W., & Molnar, L. J. (2009). Older Adult Safety and Mobility. 10.1177/1087724X09334494, 13(4), 288–300. 10.1177/1087724X09334494

Emerson, J. L., Johnson, A. M., Dawson, J. D., Uc, E. Y., Anderson, S. W., & Rizzo, M. (2012). Predictors of driving outcomes in advancing age. Psychology and Aging, 27(3), 550–559. 10.1037/A0026359

Fraade-Blanar, L. A., Ebel, B. E., Larson, E. B., Sears, J. M., Thompson, H. J., Chan, K. C. G., & Crane, P. K. (2018). Cognitive Decline and Older Driver Crash Risk. Journal of the American Geriatrics Society, 66(6), 1075. 10.1111/JGS.15378

Hamano, T., Takeda, M., Sundquist, K., & Nabika, T. (2016). Geographic Elevation, Car Driving, and Depression among Elderly Residents in Rural Areas: The Shimane CoHRE Study. International Journal of Environmental Research and Public Health, 13(5). 10.3390/IJERPH13070738

Hanson, T. R., & Hildebrand, E. D. (2011). Are rural older drivers subject to low-mileage bias? Accident; Analysis and Prevention, 43(5), 1872–1877. 10.1016/J.AAP.2011.04.028

Jo, O., Kruger, E., & Tennant, M. (2021). Public transport access to NHS dental care in Great Britain. British Dental Journal 2021, 1–9. 10.1038/s41415-021-3002-3

Kosuge, R., Okamura, K., Kihira, M., Nakano, Y., & Fujita, G. (2017). Predictors of driving outcomes including both crash involvement and driving cessation in a prospective study of Japanese older drivers. Accident Analysis & Prevention, 106, 131–140. 10.1016/J.AAP.2017.05.019

McGwin, G., Owsley, C., & Ball, K. (1998). Identifying crash involvement among older drivers: agreement between self-report and state records. Accident; Analysis and Prevention, 30(6), 781–791. 10.1016/S0001-4575(98)00031-1

Merlin, L. A., Guerra, E., & Dumbaugh, E. (2020). Crash risk, crash exposure, and the built environment: A conceptual review. Accident Analysis & Prevention, 134, 105244. 10.1016/J.AAP.2019.07.020

Morrissey, S., Jeffs, S., Gillings, R., Khondoker, M., Patel, M., Fisher-Morris, M., Manley, E., & Hornberger, M. (2024). The Impact of Spatial Orientation Changes on Driving Behavior in Healthy Aging. *The Journals of Gerontology. Series B*, Psychological Sciences and Social Sciences, 79(3). 10.1093/GERONB/GBAD188

Morrissey, S., Jeffs, S., Gillings, R., Khondoker, M., Varshney, A., Fisher-Morris, M., Manley, E., & Hornberger, M. (2024). GPS navigation assistance improves driving mobility in older drivers. 10.31234/OSF.IO/ZD79J

O’Connor, M. L., Edwards, J. D., Small, B. J., & Andel, R. (2012). Patterns of Level and Change in Self-Reported Driving Behaviors Among Older Adults: Who Self-Regulates? The Journals of Gerontology Series B: Psychological Sciences and Social Sciences, 67(4), 437. 10.1093/GERONB/GBR122

Office for National Statistics. (2024). Key Findings, Statistical Digest of Rural England - GOV.UK. https://www.gov.uk/government/statistics/key-findings-statistical-digest-of-rural-england/key-findings-statistical-digest-of-rural-england

Oxley, J., Langford, J., & Charlton, J. (2010). The safe mobility of older drivers: a challenge for urban road designers. Journal of Transport Geography, 18(5), 642–648. 10.1016/J.JTRANGEO.2010.04.005

Payyanadan, R. P., Lee, J. D., & Grepo, L. C. (2018). Challenges for Older Drivers in Urban, Suburban, and Rural Settings. Geriatrics, 3(2). 10.3390/GERIATRICS3020014

Pucher, J., & Renne, J. L. (2005). Rural mobility and mode choice: Evidence from the 2001 National Household Travel Survey. Transportation, 32(2), 165–186. 10.1007/S11116-004-5508-3/METRICS

Ritter, A. S., Straight, A., & Evans, E. (2002). Understanding Senior Transportation: Report and Analysis of a Survey of Consumers Age 50+. *Washington*, *DC*: *AARP Public Policy Institute*.

Stefanidis, K. B., Mieran, T., Schiemer, C., Freeman, J., Truelove, V., & Summers, M. J. (2023). Cognitive correlates of reduced driving performance in healthy older adults: A meta-analytic review. Accident Analysis & Prevention, 193, 107337. 10.1016/J.AAP.2023.107337

Strogatz, D., Mielenz, T. J., Johnson, A. K., Baker, I. R., Robinson, M., Mebust, S. P., Andrews, H. F., Betz, M. E., Eby, D. W., Johnson, R. M., Jones, V. C., Leu, C. S., Molnar, L. J., Rebok, G. W., & Li, G. (2020). Importance of Driving and Potential Impact of Driving Cessation for Rural and Urban Older Adults. The Journal of Rural Health, 36(1), 88–93. 10.1111/JRH.12369

Thompson, J. P., Baldock, M. R. J., Mathias, J. L., & Wundersitz, L. N. (2013). An examination of the environmental, driver and vehicle factors associated with the serious and fatal crashes of older rural drivers. Accident; Analysis and Prevention, 50, 768–775. 10.1016/J.AAP.2012.06.028

Vivoda, J. M., Heeringa, S. G., Schulz, A. J., Grengs, J., & Connell, C. M. (2017). The Influence of the Transportation Environment on Driving Reduction and Cessation. The Gerontologist, 57(5), 824. 10.1093/GERONT/GNW088

Wang, C., Sierra Huertas, D., Rowe, J. W., Finkelstein, R., Carstensen, L. L., & Jackson, R. B. (2021). Rethinking the urban physical environment for century-long lives: from age-friendly to longevity-ready cities. Nature Aging 2021 1:12, 1(12), 1088–1095. 10.1038/s43587-021-00140-5

Zwerling, C., Peek-Asa, C., Whitten, P. S., Choi, S. W., Sprince, N. L., & Jones, M. P. (2005). Fatal motor vehicle crashes in rural and urban areas: decomposing rates into contributing factors. Injury Prevention, 11(1), 24–28. 10.1136/IP.2004.005959

