## Supplementary Information file for "The impact of urban vs rural environments on driving in ageing"

### **Supplementary Section**

| **Supplementary Table 1. Driving mobility and safety measure descriptions** | | |
| --- | --- | --- |
| Measure: | Questionnaire: | Description: |
| Annual mileage | Driving History | Participants were asked “What is your annual mileage in a typical year?” |
| Driving days | DHQ | Participants were asked the average number of days driven per week (ranging from 0 to 7). |
| Driving space | DHQ | Participants were asked how often they drive within 6 geographical areas, from within their immediate neighbourhood (lowest), to outside their region (highest). For each question, scores were rated from one (a few times in the year) to four (every day). Totalled scores across all six items comprised driving space. |
| Weekly trips | DHQ | Participants were asked “How many trips per week?” for each location they typically drive to. Totalled scores comprised weekly trips. |
| Maximum weekly trip distance | DHQ | Participants were asked to provide the “Estimated miles from home (single trip, one-way)” for each location they typically visit per week. The maximum single-trip distance comprised maximum weekly trip distance. |
| Situational avoidance | DHQ | Participants were asked whether they completed a particular challenging driving situation within the past 3 months (i.e., driving in the rain). The totalled number of situations avoided per participant comprised a situational avoidance measure, ranging from nought to eight. |
| Relative driving speed | DHQ | Participants were asked “How fast do you usually drive compared to the general flow of traffic?” and rated their answer on a five-point Likert scale (Much slower – Much faster). |
| Transport preference | DHQ | Participants were asked “which way do you prefer to get around?” and selected one of “Drive yourself/ Have someone drive you/ Use public transportation or a taxi”. |
| Recent road incidents | RTI | Participants were asked how many RTIs they experienced in their driving history, and when their most recent RTI was. A recent RTI was classed as an RTI taking place within 3 years of data collection (since 2018). |

| **Supplementary Table 2.** **Linear mixed effect model analysis showing how rural and urban environments influence driving mobility over time** | | | |
| --- | --- | --- | --- |
| **Variable** | ***B*** | **t-value** | ***p*** |
| Driving space | -0.65 | -2.88 | <.01 |
| Driving days | -0.11 | -1.06 | ns. |
| Max. trip distance | -2.04 | -1.13 | ns. |
| Weekly trips | -0.49 | -1.56 | ns. |
| Situational avoidance | 0.33 | 3.34 | <.001 |
| Driving speed | 0.03 | 0.68 | ns. |
| *Note*.  ^a^ Displaying unstandardised beta coefficients | | | |
